# Intersectional Experiences of Non-Communicable Diseases and Health Seeking Strategies in Informal Settlements in Freetown, Sierra Leone

**DOI:** 10.1101/2025.10.03.25336572

**Authors:** Abu Conteh, Laura Dean, Annie Wilkinson, Joseph Macarthy, Braima Koroma, Sally Theobald

## Abstract

This paper explores the burdens of non-communicable diseases (NCDs) in low-income settings, and how they are shaped by structural barriers including gender inequities. As Low- and Middle-Income Countries (LMICs) face epidemiological transitions towards increased NCD burdens, the challenges faced by people living with NCDs are a priority. We employed narrative inquiries to explore the lived experiences and healthcare seeking decision making practices of 15 participants living with diabetes, hypertension, and disability related to stroke in three informal settlements in Freetown. Narrative interviews were conducted through three household visits within a period of 12 weeks. We adapted the Intersectional Gender Analysis Framework for Infectious Diseases of Poverty Research to understand how poverty, gender and other axes of inequity interact with NCD burdens in informal settlements. Findings show a strong connection between poverty, gender identities and comorbidities linked to NCDs. Women’s lived experiences of NCD conditions reflected historical disadvantage and patriarchal oppression, most notably through their limited financial autonomy, barriers to healthcare decision making and treatment access, compounded by gendered impacts of conflict and migration. Men’s experiences were however influenced by changes in social status, due to conflict and migration, and financial instability, limiting access to healthcare. Gender differences were also key in shaping household and healthcare decision making, as gender norms and experiences of masculinities and femininities reflected the division of roles and access to resources by men and women, which in turn shaped their ability to seek early and better healthcare interventions. In conclusion, our study has shown that for people living with NCDs, gender norms and patriarchal structures reinforce power hierarchies, worsen health outcomes and deepen poverty. Healthcare interventions must consider the full range of needs and impacts of people impacted by long term illnesses and the context in which they live.

## Introduction

Globally, NCDs account for an annual death rate of about 41 million (Yuyun et al., 2020; Mahumud et al., 2022), 77% of which affect LMIC settings (WHO, 2022). Among the growing burden of NCD conditions, cardiovascular diseases (CVDs) pose the greatest concern, as they cause the most deaths worldwide. WHO estimates that CVDs alone cause over 17 million deaths annually, more than three quarters of which occur in LMICs. This staggering rate of morbidity and mortality requires urgent action, as this has the potential to undermine the response capacity of health systems in these settings - as they grapple with multiple and changing health stressors and crises.

As LMICs face epidemiological transitions such as the interactions between NCD and infectious diseases, (Thienemann et al., 2020), people living with NCDs can be severely impacted, due to broader social and structural drivers such as conflict, poverty, migration, health and housing inequalities. Sierra Leone is no exception to global NCD burdens, as over 70% of the population is exposed to risk factors such as unhealthy diets, tobacco use, alcohol abuse and air pollution, which contribute to about 30,000 deaths annually (Government of Sierra Leone 2020a). Among the top ten causes of death in Sierra Leone, ischemic heart disease and stroke are ranked 4^th^ and 6^th^ respectively (Government of Sierra Leone, 2020b). This study focuses therefore on understanding the lived experiences of people living with diabetes, hypertension and disability resulting from stroke in informal settlements in Freetown, Sierra Leone, and how these experiences shape their health seeking strategies.

With the rapid rate of urbanisation, and the spread of urban informal settlements, NCDs are a new layer to existing challenges, as the rate of urban growth has outpaced the provision of services, including healthcare. In addition to service access challenges, poverty and precarious living conditions reinforce NCD vulnerabilities, particularly in informal settlements. A study conducted in an informal settlement in Freetown - Kroobay, identified diabetes and hypertension as some of the NCD conditions informal settlement residents suffer from due to poor diets, smoking and limited physical activity (Kamara et al., 2023). Social and environmental risk factors e.g. smoke pollution, financial stress, poor access to housing, clean drinking water and healthy food are commonplace in informal settlements in Freetown (Wilkinson et al., 2020). While the structural barriers shaping poor health outcomes are being acknowledged within policy, mechanisms for addressing these barriers tend to be fragmented, and impacted by gaps in evidence. NCDs, interacting with poverty and social inequality can therefore be particularly detrimental to people living in informal settlements. As highlighted by Elsey et al., (2019) addressing systemic urban health inequalities, particularly in the face of rising NCD threats requires a rethink of how the health system should work to address the broader social determinants of health and include the voices of the urban poor in decision-making.

Within the vulnerable populations in informal settlements, a further axis of disadvantage is gender. The creation of gender-blind health policies and interventions in fragile settings signals that the healthcare needs of women and men affected by NCDs are not met (Percival et al., 2018). Morgan et al., (2022) noted that gender-blind health policies and interventions have wider consequences beyond health, citing that the lack of such considerations during the COVID-19 response negatively impacted the wellbeing of vulnerable women and men in marginalised urban settings across different contexts (Morgan et al., 2022). Gender gaps need addressing in the planning and delivery of healthcare as gender roles and norms shape how women and men experience and live with health crisis, through access to resources, and health seeking decision making (Connolly et al., 2020). However, significant gaps still exist in understanding these intersectional drivers of NCD and the diverse experiences of women and men in Sierra Leone.

### Gender, Health and other Axes of Inequity in Sierra Leone

Gender disparities in Sierra Leone underlie women’s barriers to healthcare, educational and livelihood opportunities. National statistics show that women are significantly disadvantaged when it comes to educational attainment, access to healthcare, financial services and the labour market. For example, literacy rates among women (43%) are far lower than men (62%). Additionally, gender disparities in wage earnings are huge as only 45% of employed women receive financial reward for their work, compared to 72% of men (Statistics Sierra Leone, 2019). These structural barriers reflect women’s lack of autonomy in household and healthcare decision making, including sexual and reproductive health rights (Action Aid Sierra Leone, 2018). A gender and intersectional analysis is vital in understanding health inequalities and in pushing for an inclusive and integrated healthcare delivery system particularly for marginalised urban residents.

Our previous research into health in informal settlements indicate that gender roles, with respect to the management of long-term illness and health seeking are reinforced by social and cultural norms (Wilkinson, et al., 2020). Societal expectations of women firmly place caregiving roles into their hands, while decision making rights (including healthcare) are accorded to men, which negatively impact women’s health outcomes (Conteh et al., 2021). A study in a Freetown informal settlement found that women’s uptake of COVID-19 vaccines and related healthcare services, (e.g. reproductive health services) were hindered by livelihood, and care responsibilities, influenced by historical barriers to education and the lack of inclusion in decision making (Conteh et al., 2023). These experiences of access to preventive medicine including COVID-19 vaccines were different for men as they were more likely to be formally employed which created more incentive for them to be vaccinated through regulations on the mandatory possession of vaccine cards to access public buildings (Conteh et al., 2023).

While these gendered health outcomes exist, it has been established that men also face health crisis. Men’s experiences of health crisis are shaped, not least because of physical manifestations of disease, but due to the loss of financial autonomy and respect (Vlassoff, 2007). While these studies focused on the intersections of gender and health, they did not explicitly focus on gender asymmetries as a driver of NCDs outcomes.

This study therefore builds on our current understanding of gender hierarchies and their interactions with broader structural drivers such as poverty in shaping the experiences of NCD outcomes and health seeking strategies. An intersectional analysis approach was applied, informed by the Intersectional Gender Analysis Framework for Infectious Diseases of Poverty Research (WHO 2020) to explore these relationships through patients’ perspectives in informal settlements in Freetown. The study explores health seeking drivers and the role of informal healthcare providers (alongside biomedical providers) in the provision of NCD care in the context of healthcare access challenges faced by urban informal settlement residents.

## Materials and Methods

### Study setting and link within broader ARISE research programme

This study was conducted in three informal settlements in Freetown which face challenges with service delivery, including access to water, sanitation, and healthcare. Our research sites included Cockle Bay, which is a seaside settlement in the west of Freetown, and Dwarzark and Moyiba which are both hillside settlements in the central and far east of the city. The settlements were purposively selected to ensure diverse voices in informal settlements, and exploration of how their experiences about the places they live in shape health experiences and treatment seeking. Our approach was in recognition of the need to understand the peculiarities of informal settlements, despite the similar challenges they face, which is important to the design of health programmes particularly for people with long term illnesses. The study was undertaken within the Accountability, Responsiveness and Equity Hub (ARISE) which worked with informal settlement residents. including co-researchers (people living and working in informal settlements) in addressing intractable health challenges (ARISE, 2024). This working relationship aimed at strengthening equitable partnerships with communities (Ozano et al., 2024), and in improving health and wellbeing in marginalised settings in Freetown.

### Methodology, methods and participants

We conducted narrative interviews to explore the lived experiences of NCD health conditions and health seeking priorities. The interviews were conducted through three household visits to a cohort of 15 participants living with diabetes, hypertension, and disability resulting stroke within a period of 12 weeks. Narrative interviews form a part of the narrative inquiry methodology which allows participants to reflect on their lived experiences, within broader social, political and environmental contexts (Creswell, 2007). We explored participants’ personal life circumstances including family history, barriers to education, livelihood attainment and their interactions with health outcomes. The interviews involved discussing participants’ visual diaries of the different healthcare providers they visited (formal and informal providers) in the weeks prior to each household visit and a reflection on the choice of preferred providers, the type of conditions presented and the outcomes of the treatments they received. The visual diaries included animated objects such as a health centre, a herb and a herbalist, representing the different providers to help participants to recall their healthcare visits.

### Recruitment of participants

We designed a purposive sampling strategy to identify and recruit participants for the narrative interviews. Participant selection was based on the type of NCD conditions and other identities including age, gender, and place of living within the settlements to achieve a diversity of participants’ characteristics and perspectives (Nyimbili, 2024). Selection was informed by the pre-determined criteria listed above. Participants were informed about the study and asked whether they wanted to participate. The participants were identified by co-researchers (community members participating in research) based on agreed criteria. Those who met the selection criteria were selected in consultation with co-researchers and interview dates agreed.

The recruitment of participants for this study was done from 6^th^ to 10^th^ September 2021, while the three interview sessions were held from 13^th^ September to 14^th^ December 2021.

Participation in the study was based on informed consent for which participants were provided with adequate information to allow them to decide their participation. Participants were informed about their rights in the study including their right to withdraw at any time, without reprisals. The rights of participants also included seeking clarifications or requesting for interviews to be paused if they felt uncomfortable about sensitive topics. Before the start of each interview session, participants were asked to sign a written consent form after being provided with information about the consent procedures. Table 1 below describes the characteristics of participants whose names were pseudonymised to conceal their identity.

**Table 1:**
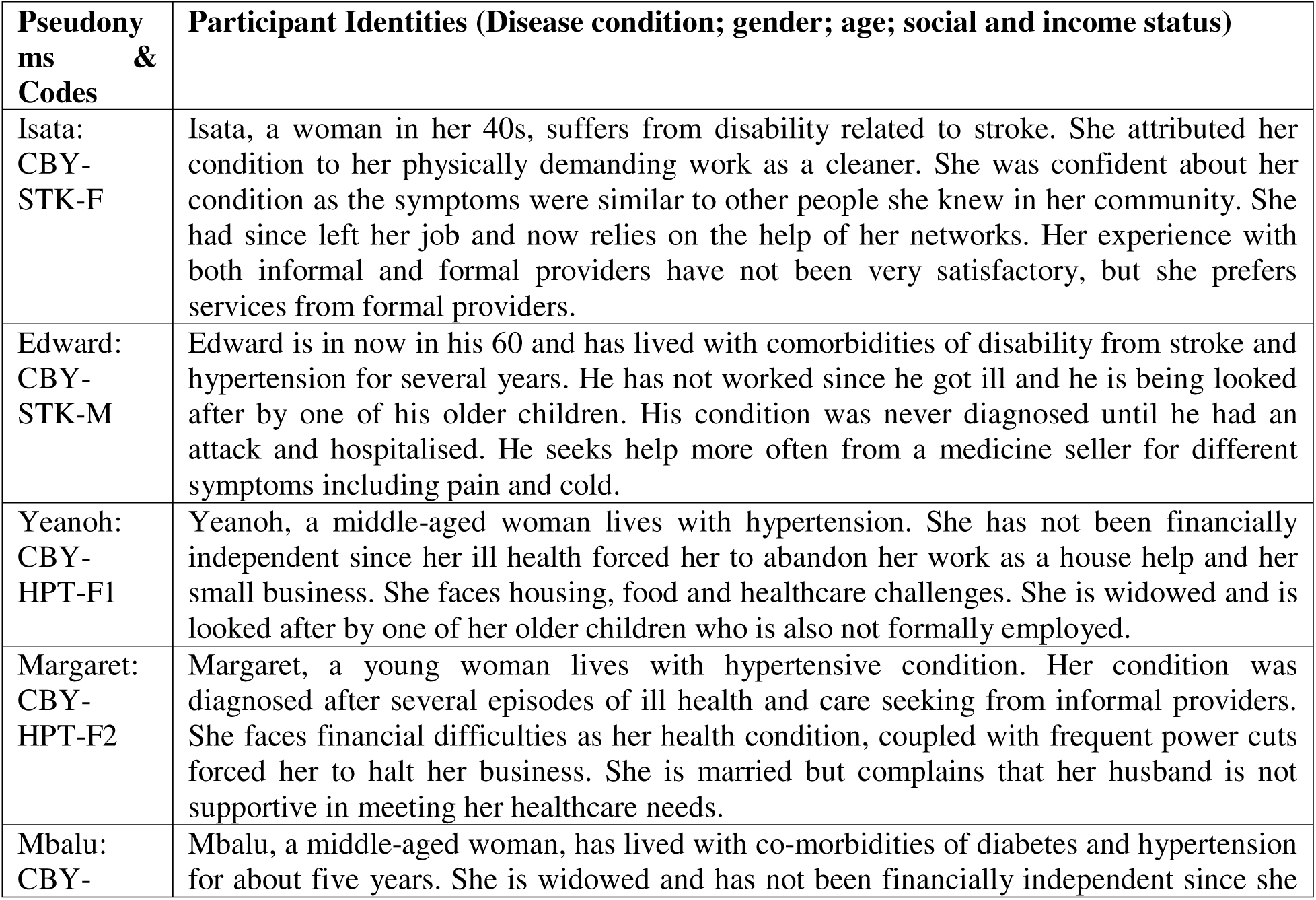
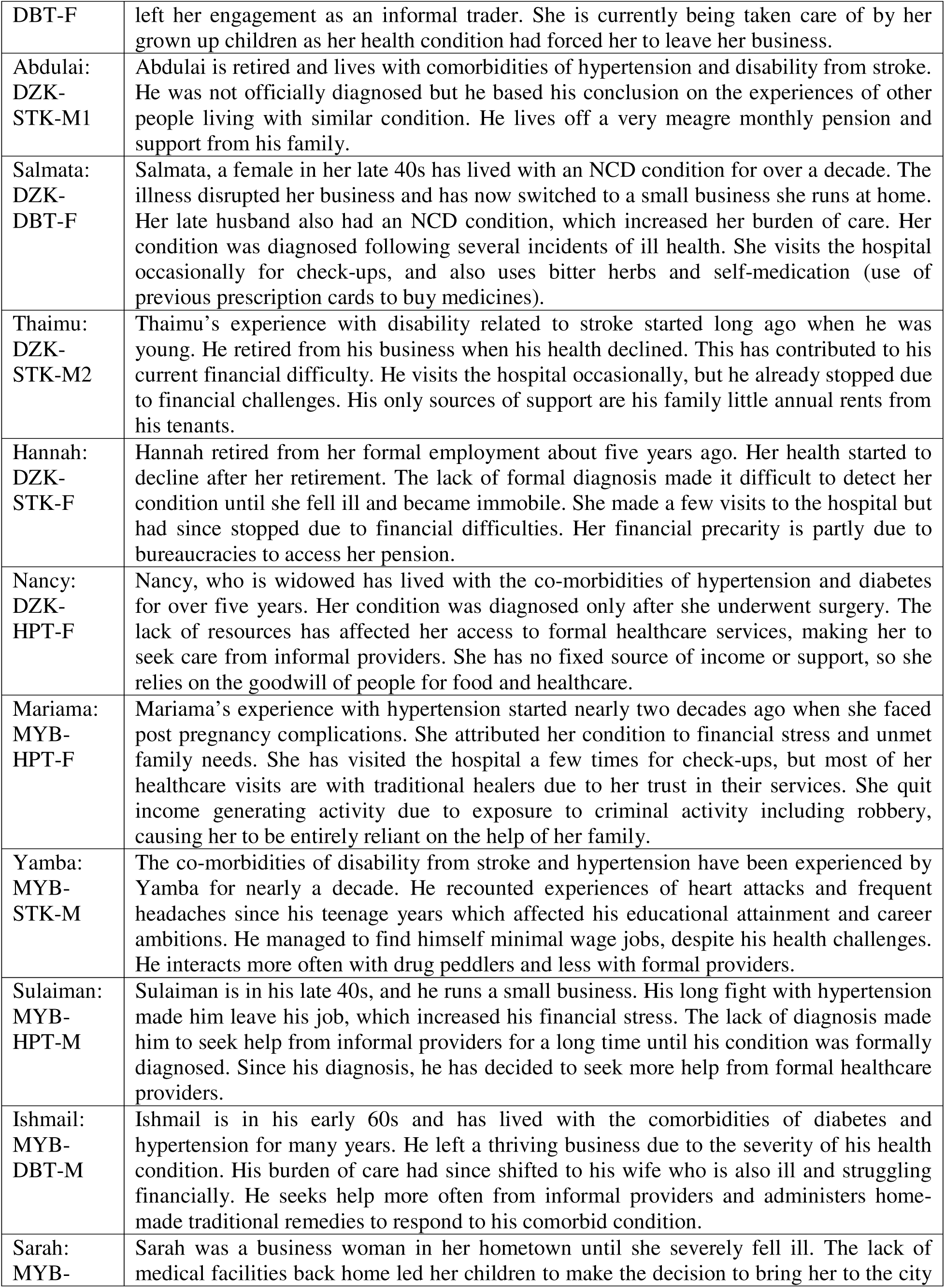
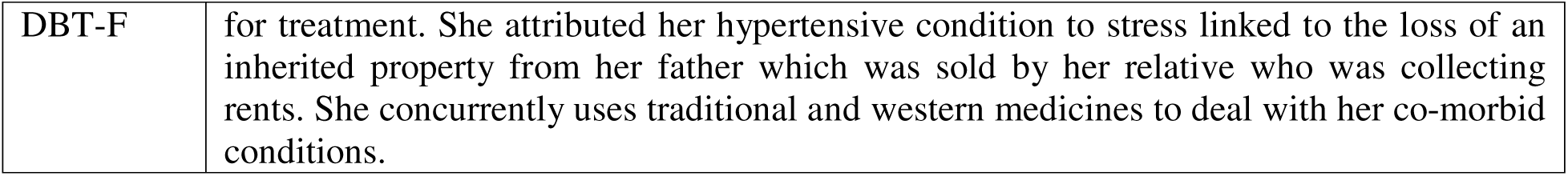
Description of Narrative Interview Participants.

### Analysis

This study took an intersectional analysis approach informed by the Intersectional Gender Analysis Framework for Infectious Diseases of Poverty Research (WHO, 2020) which provides extensive guidance on how to generate, analyse and interpret findings on disease interactions with poverty, gender and other axes of inequity. The framework was applied in an urban informal settlement context in Freetown, to understand the complex structural drivers shaping NCD vulnerabilities (Kamara et al., 2023), mediated by poverty and precarious living conditions (Wilkinson et al., 2020). Our adaptation of the infectious disease framework reflects the need to approach NCD policy from a social perspective, beyond the current biomedicalisation of health. Our analysis deployed relational lenses to understand the gendered experiences of NCD health problems. These experiences were reflected upon throughout our analysis as they were viewed as drivers that shape people’s lived experiences of long-term health problems. This involved reading and re-reading transcripts to iteratively cluster emerging concepts. The transcripts developed from the interviews for the three visits to participants were coded separately and deductively analysed to observe the patterns in which concepts were developed into codes. The codes were then organised and clustered into similar concepts to generate themes and sub themes.

This approach was useful in understanding participants’ framing of gender norms, household power dynamics and healthcare decision making based on different identities. We reviewed and adapted the Intersectional Gender Analysis Framework for Infectious Diseases of Poverty Research as it aligned to our themes (See Figure 1 below). Findings from the themes were first summarised into a table articulating their linkages and contribution to the study objectives and the conceptual framework. The texts from the tables were then applied to guide the adaptation of the conceptual framework and to clearly articulate the study findings.

**Figure 1:**
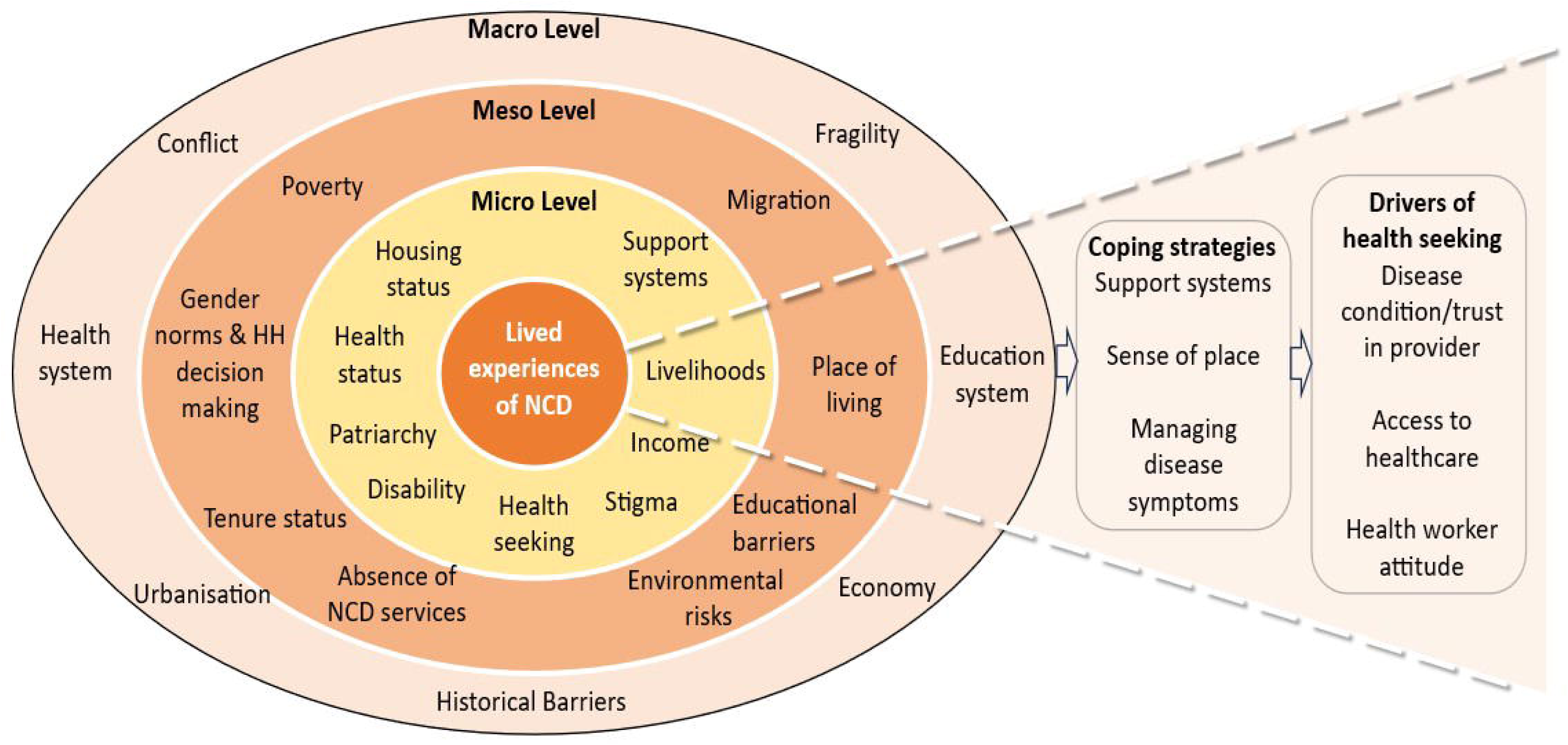
Intersectional Gender Analysis Framework for Research on NCDs (Adapted from WHO’s Intersectional Gender Analysis Framework for Infectious Diseases of Poverty (WHO 2020)

### Ethics and safeguarding

Ethics approvals were sought and granted by the Sierra Leone Ethics and Scientific Review Committee on 3^rd^ of August 2021 (Version 2.0 of 28 July 2021). Approval was also granted by the Research and Ethics Committee of the Liverpool School of Tropical Medicine on 5^th^ August 2021 with reference number 21-043. The research team discussed during the debrief sessions adhering to ethical procedures by ensuring that research participants and vulnerable adults living with NCD conditions were protected from harm alongside the research team. The research team and participants were informed about referral procedures in the event of safeguarding concerns such as sexual exploitation and abuse of women and children or research participants. The phone contact of the Safeguarding Lead for this study was made available to all participants through the information sheets that were provided. Participants who faced distress during the interviews were allowed to grieve and were consoled. Requests were made to refer the participants to professional counselling services. However, no referrals were made as participants reported that they were relieved after discussing their challenges with the first author (AC).

Data processing and analysis were done from January to August 2022. Data was then made available to authors for review and in planning the writing of this manuscript. To enhance confidentiality and data protection, adequate safeguards were made to protect the identities of participants in accordance with the data protection procedures of LSTM and SLESRC. This included the deidentification of participants and labeling transcripts with unique identification codes before sharing them with authors for review. The coding framework, and an outline of the manuscript were shared with all authors through email which informed the discussions for the writing of this manuscript. All transcripts and analysed data were stored on Nextcloud, a secure data repository hosted by LSTM. Access to the database was granted only to the first author, AC and supervisors of this study based at LSTM, ST and LD.

## Results

Findings from this study are presented in three parts. First, we explore participants’ lived experiences of NCDs, and how they are mediated by gender norms, access to resources and participation in decision making. Second, we describe participants’ coping strategies and the way these are shaped by personal circumstances such as gender, disability, co-morbidities, disease severity, and tenure status. Third, we describe the drivers of health seeking, shaped by health system challenges and treatment outcomes. Figure 1 below reflects the key findings presented in the three sections (lived experiences, coping strategies and drivers of health seeking) showing how these are shaped by three layers of interrelated forces (macro, meso and micro). The macro factors represent the structural drivers at city/national level, meso factors at the community level, and the micro factors at individual or household level. Table 2 (at the bottom of the text) also aids the interpretation of the in-text codes.

**Table 2:** Description of In-text Codes by Alphabetical Order. This table has been provided to ease the interpretation of the codes with the text in alphabetical order

| Code name | Meaning | Code name | Interpretation |
| --- | --- | --- | --- |
| CBY | Cockle Bay | M | Male |
| CC | Community chief | MYB | Moyiba |
| DBT | Diabetes | NI | Narrative Interviews |
| DP | Drug peddler | RH | Religious healers |
| DZK | Dwarzark | STK | Stroke |
| F | Female | TH | Traditional healer |
| HPT | Hypertension |  |  |

### Lived Experiences of NCDs

Lived experiences of NCD reflected the connections between gender norms, structural barriers and their contributions to social and health outcomes and healthcare decision making, as evident in our findings. We demonstrate that gender and societal expectations about men and women reinforce power hierarchies in decision making, leading to worsening health outcomes and poverty. Figure 1 above summarises our adaptation of WHO’s Intersectional Gender Analysis Framework, applied in our analysis to explore the relationships between poverty and NCD outcomes, and how these influence participants’ response to NCDs and health seeking strategies. The framework reflects the interactions between the intersectional drivers of NCD outcomes and broader social and structural factors that shape how people living with NCDs cope with their conditions and seek healthcare. To illustrate these interactions, we present the stories of Isata in box 1 and Ishmail in box 2 with the aim of providing a fuller and illustrative understanding of the lived experiences of people living with chronic disease.

#### Box 1

**Isata’s Story**

Isata lives with her family in a rented single bedroom apartment built with corrugated iron sheets or “pan-body”. She lives with comorbid conditions of hypertension and disability related to stroke, which she relate to mental stress, depression and precarious working condition. She used to work as a cleaner which involved strenuous activities, including the use of water for a long period of the day. Although she did not have a formal diagnosis, she was convinced that she had developed stroke: ‘‘*I lost strength in my arms, my feet became numb, with restricted movement - some form of disability…… I observed some of the symptoms from people with stroke. My friend lives with stroke.”*

Isata got married at a relatively young age after she had dropped out from school. She had a rough childhood because of what she attributed to neglect by her father, and the poor health condition of her mother following her birth, leaving her in the care of a family caregiver. She said that the lack of support from her father and the precarious financial situation of her caregiver deprived her of education: *“My father did not care for me…. So, I stopped at class five. So, I do not consider that as education”*.

Her relationship with her partner turned sour because of his alleged alcoholic and abusive behaviour, and the neglect of his financial responsibilities. Isata reflected on her continued stress over the high burden of childcare with little income from her previous menial job, meaning, she had to rely on the support of others. *“I have been the one taking care of my children and myself. One of them is in the University, and the burden is all on me”*.

Isata’s financial uncertainties due to her ill health and the loss of livelihood options means limited access to healthcare. The lack of resources has led her to try different healthcare options including traditional healers. Her initial experience with formal healthcare providers was not favourable because of the lack of clear diagnosis, thus increasing her stress about her recovery. Despite this, she prefers seeking care from formal healthcare providers whenever she has money because of the relatively better outcomes, compared to traditional healers. The outcome from her hospital visits, she said was helpful. She combined these interventions with spiritual healing and prayer sessions: “W*ith God’s intervention, the pain was fading away, and Therefore, the combined forces made the difference”*.

#### Box 2

**Ishmail’s story**

Poverty and migration from rural areas are the common experiences of people living in informal settlements in Freetown. This is reflected in the accounts of Ishmail who left his native rural village to look for a better life. The death of his parents cost him the opportunity of formal education. His business adventure was successful, from which he built himself a house.

Soon, Ishmail’s fortune would be hijacked by an uncertain future –long term ill health. He has lived with the co-morbidities of diabetes and hypertension for many years. He explains that his health condition has plunged him into deep financial crisis since he was diagnosed. This means that he had to rely on his family members (some of whom are unemployed) for support. Ishmail’s account reflects the shrinking support system for the chronically ill as their carers also face financial and emotional crisis: *“My wife is currently not able to do business; we hugely depend on our children but even our children do not have good job….”.* He feels embarrassed that he is being taken care of by his wife.

His usual experience with hypertension is headache, fatigue, perspiration and blurry vision. For diabetes, he explains that although he doesn’t have the machine to check his blood sugar, he experiences itchiness whenever his blood sugar increases.

Ishmail has had limited interaction with the formal health system, partly because of financial limitations and his health belief. While he believes in the efficacy of western medicine (he occasionally visits the hospital for checkups when he has money), he also trusts traditional remedies made from herbs such as mango leaves, guava leaves and bitter roots for the treatment of diabetes and hypertension: “I am not saying medicines from the formal health service providers are not good…. *but the herbs are effective for diabetes and hypertension, and I always see massive improvement”*.

### The interplay between gender and poverty: barriers to education

The health outcomes and reduced capacity to respond to the health crisis for men and women is often similar. However, gendered hierarchies and the influence of other factors such as minimal education often meant that the route to this outcome was different for men and women. Throughout our findings, women expressed difficulties in accessing educational resources as in the case of Isata (box 1), which limited their capacity to respond to health crisis. For example, women living with co-morbid diabetes and hypertension were particularly concerned about their exclusion from formal education as a result of patriarchal family values having had long-term impacts beyond their healthcare needs, affecting upward social mobility and livelihood opportunities.

Older women with co-morbidities of diabetes and hypertension, were vocal about family and patriarchal notions of women and the institution of marriage which encouraged families to give off their daughters to marriage when they were very young, with economic stability often described as a key driver. This was the case with Sarah:

> *“I got married to a man who had two wives already; I was the third. He was the same man who took me from school. I am the last child of my mother and when she died there was no money to further my education, so I was forced to marry this man.” (NI_MYB_DBT_F)*

> *“The reason why I did not go to school was because my father valued male education more than female education, despite people telling him about the benefits of girlchild education… I feel extremely bad that I did not receive formal education. My life would have been much better today if I had formal education.” (NI_DZK_HPT_F).*

Care responsibilities were also reported to have unduly affected women’s educational advancement, which also impacted their future career prospects. One woman living with co-morbidity of diabetes and hypertension *(NI_CBY_DBT_F)* described that her role as the prime caregiver of the child of her deceased elder sister took a toll on her schooling and eventually led to her withdrawal from school.

Men also shared their experiences about the challenges of accessing formal education. However, these experiences seemed to be tied largely to cycles of poverty, and personal health circumstances, rather than driven by patriarchal disadvantage as in Ishmail’s story (Box 2). For example, some men described that the loss of a breadwinner during childhood meant assuming family responsibility at an early age, disrupting the chances of career progression. A boy having to take early family responsibility was driven by patriarchal expectations of men starting from the early years of life:

> *“My parents died, and I was left with my relatives who also had their own families to take care of. I didn’t want to overburden them, so I came to the city. Thankfully, I learned a technical skill from which I supported my family back home.” (NI_DZK_HPT_M)*

Men also stated that personal health conditions acted as barriers to educational access and completion. Many adults in Sierra Leone engage in schooling to attain basic literacy, in response to lost opportunities to be educated during their formative years. An elderly man with stroke and hypertensive conditions, noted that he faced strange health problems while in school, climaxing to several heart attacks and headaches forcing him to leave school early:

> *“I left school because of this illness. I think I had the first attack when I was preparing to complete my school leaving examination, but the agonising headaches forced me to discontinue the preparations for the exams.” (NI_MYB_STK_M).*

### Financial hardship: The interplay between gender, poverty, ill-health and fragility

Experience of financial hardship was common for participants, and this was due to both non-health and health factors. First, as an outcome of social and political instability such as the civil conflict, forced migration and the lack of livelihood opportunities. Second, as an outcome of chronic long term health problems.

Participants stated that the civil conflict forced migration, and the loss of livelihood opportunities severely impacted their financial status. Participants migrating to urban settings expressed that they had high hopes of better living conditions, but their hopes were shattered due to the new layers of vulnerabilities faced in urban informal settlements, including limited housing access, poor housing conditions, and insecure livelihoods. For some people affected by NCDs, living in an informal settlement brought about new challenges which contributed to anxiety, and stress. Men and women in these conditions spoke about difficulties faced to secure a decent living due to livelihood disruptions. However, some men found themselves lucky enough to establish and sustain small businesses, whilst others felt less fortunate, undertaking menial jobs such as night security, laundry, and other forms of domestic labour as an immediate solution but with ongoing financial insecurity. Older generations of migrants, who arrived in Freetown before the war, also spoke about the impacts of the war and military coups which left them unemployed. Some men described that the long-term effects of conflict and insecurity made them more vulnerable to the poverty-illness nexus.

> *“I became jobless since the military coup in 1992, and I have not been able to get a job ever since. To survive and sustain my family, I engaged in menial jobs like ironing, laundry, and other domestic chores.” (NI_CBY_STK_M_02)*

> *"We suffered a lot during the civil war. After I lost my job at the inception of the war, I decided to go into a small business, but the rebels destroyed everything when they attacked our town. I was left with nothing. When I came to Freetown, life was difficult because I knew no one and I had no job which got me frustrated.” (NI_MYB_HPT_M)*

Women’s accounts of poverty and financial difficulties were centred on historical experiences. Food and housing difficulties, and personal circumstances such as widowhood, and single parenting also drove financial instability. Irrespective of whether women had been born in Freetown or migrated to informal settlements later in life, they experienced similar financial difficulties which they believed were negatively impacting their quality of life.

For some women, they had engaged in precarious livelihood activities due to financial challenges within their families, which has exposed new layers of vulnerabilities as described by Mariama *(NI_MYB_HPT_F):*

> *“I live with my husband who is jobless; this difficult situation left me with no choice but to go into wood harvesting and selling to survive with my family……I left wood selling due to fear of armed robbers and kidnappers in the bush. I also engaged in stone quarrying for four years until when I gave birth to twins and later developed health complications.” (NI_MYB_HPT_F)*

Absence of family to support with childcare where women had become widowed during the war following rural-urban migration also led to gendered challenges.

> *"When we came to Freetown, it was extremely difficult for us. I had engaged in buying “Mina fish” (small sized and cheap fish) at the beach and hawked across several neighbourhoods to make a living. I discontinued fish hawking and started selling cake which I continued till we moved into this community.” (NI_CBY_DBT_F)*

Poverty and financial instability were often exacerbated as a result of NCDs, further disrupting livelihoods and pushing people further into the poverty-illness nexus. Women and men stated that because of their recurring health problems financial pressures were mounting due to their difficulty in sustaining livelihood activities and additional costs of health seeking (discussed later). Despite these shared financial difficulties for both men and women, outcomes were different related to the severity of health problem, alternate income and support systems.

Salmata (*NI_DZK_DBT_F)*, a middle-aged woman living with diabetes (who was also diabetic) narrated her experience with ill health which she said had caused her to abandon her food retail business in the market:

> *“I quit doing business at the market because of diabetes. My health condition was seriously affecting my ability to do business.” (NI_DZK_DBT_F)*

Experiences of chronic long-term ill health and their impact on livelihood outcomes were described by some women as causing them to lose all hopes of a better life.

> *"Honestly, I have no plans because there is nothing, I can do to generate income. I have been in this condition for over a year now and my health is not improving… ” (NI2_CBY_STK_F)*

Within gendered categories, differences in experience were also evident. For example, men who were low-income earners and with high numbers of dependents were more likely to be impacted by financial hardship than men who were taken care of by their families. A male participant living with hypertension *(Sulaiman:_NI_MYB_HPT_M)* stated that having to leave his job due to poor health condition meant huge financial consequences:

> *“The reason why I resigned from my job was because I could no longer work and look for medication at the same time. This is my fourth year since I became ill and during this period, I have not been able to find a living.” (NI_MYB_HPT_M).*

### Gender Norms and Intra-Household Decision Making

Gender was identified as an underpinning power dynamic within households, shaping access to and control of available resources, ultimately shaping how people respond to disease conditions. Findings here reflect these dynamics, as they relate to masculinity, conflict and changing household dynamics, and decision making.

Masculinity and gender dynamics were seen as important in the way households shared responsibilities. Across settlements, stereotypical gender roles with women expected to nurture children while men serving as breadwinners were evident. Masculinity was often perceived in terms men’s physical and financial powers. Thus, when men became unemployed or financially vulnerable due to long term ill health, their masculinity was challenged as this meant a diminished role within the home. Many men felt pressured to keep up with these economic obligations, as these often contributed to marital conflicts, including denial of sex from their spouse which further challenged masculinities. One woman remarked that because women are always expected to be prepared for sex with their partners refusing to provide it can be a cause of conflict. In her case, sexual agency tended to be used as a strategic resource to seek her husband’s support towards meeting her healthcare and related needs:

> *“Sometimes, I tell him the fact that he is not doing the right thing because when you are in a relationship with someone, it is your responsibility to care for your partner, especially regarding health issues. He often talks about extended family burden, but I always tell him that I am his immediate family. So sometimes I deny him sex which often results in conflict.” (NI_CBY_HPT_F_02)*

For women affected by NCD conditions, where they felt that their partner was not working hard enough to meet their financial needs, they experienced an additional burden, as they were also expected to support household income.

Some women ascribed this additional burden of income generation to their ill-health. For example, Isata (Box 1) who had experienced a disability resulting from stroke attributed her condition to her precarious working conditions, combined with the attitude of her husband whom she said was not supportive:

> *“My husband does not strive hard to care for me and our children. Hence, I had to engage in menial jobs. That is what I have been doing to make ends meet which may have resulted in this illness.” (NI_CBY_STK_F).*

The intersections of masculinity, perceived loss of power, ill health and financial vulnerability were key to how men viewed themselves. Participants revealed that it was often difficult to negotiate power between men and women particularly when men felt a sense of vulnerability to ill health. In such situations, conflict was the norm, which was described as one of the crucial factors shaping how men cope with health problems. Men were described as finding it disturbing to be looked after by their wives when they face health crisis, as this signalled the “loss of power” and a change in their social standing. This was described by Salmata *(NI_DZK_DBT_F*), saying that her husband) was not comfortable with her making the decisions on his behalf which was construed as wanting to ‘control him’:

> *“He thinks that I want to be in control of the home because I provide financial contribution, but that is not the case…. He always becomes angry; He tells me sometimes that I feel I am the man of the house which is not possible…. Because my financial contribution to the home is more than his, he always feels inferior.” (NI_DZK_DBT_F)*

### Living with Disability

Participants reflected on their experiences with NCDs including the physical manifestations of disease conditions, poverty, and social disruptions. NCD related impairments (or disability) were described in terms of major deviations from social and physical functioning, notably the disruptions in mobility and livelihood activities (as described above). These disruptions - sometimes leading to disability, were often described as sudden episodes of physical impairment attributed to shocks from heart attacks and stroke, which are often diagnosed late due to healthcare access barriers. Different accounts from participants referred to disability as “life changing events” which often plunged them into long periods of reduced physical and social functioning:

> *“It was just one night, when I was sleeping. I woke up with a sudden attack and fell on the ground. Immediately, my hands and feet became numb, and I couldn’t move. I couldn’t do anything. This sickness has really changed my life.” (NI_DZK-STK-F)*

Participants’ accounts reflected the gradual progression of pain, which advanced into physical disability, such as the loss of mobility, and eye impairment. The rugged nature of the environment in informal settlements, coupled with the lack of assistive technologies for people with diminishing health were shown as factors intersecting with material and social deprivations. As described by Isata (Box 1), these factors compromise health seeking choices and worsen health outcomes.

> *“Both of my legs are weak, and I cannot stand without holding or leaning on something. Before this time, I was able to walk alone even though at a slow pace but right now, I can only do that by holding a stick or getting someone to hold me.” (NI2_CBY_STK_F)*

Beyond the physical impairments, some people were concerned about disability and other forms of identity induced by chronic disease and the fear of stigma. Some people described these physical identities in relation to disease manifestation. This description was common with women living with diabetes and stroke, which they believed were physically draining, and contributing to the change of their physical outlooks. For women living with diabetes, the feeling of looking pale or emaciated frequently caused a sense of shame and stigma, as described by one woman:

> *“My body was rounded, but it has been depreciating since I had diabetes (NI_DZK_DBT_F).*

Decision making about property ownership and control also mirrored gendered and patriarchal attitudes which participants felt did not always favour women. As a result, women owning property explained that they felt obliged to transfer their power of attorney to a male partner or relative to protect it, which sometimes resulted in negative outcomes, including their dispossession from those properties. The dispossession of women from their property had health, social and financial consequences. As described by Sarah *(NI_MYB_DBT_F)*, she lost the house she inherited from her father to her half-brother because she gave the power of attorney him to rent and manage the property:

> *“I left everything in my brother’s care including the collection of rents for the house and the shops simply because he is a man; I thought he would handle everything perfectly only for him to sell the house without my knowledge. My brother connived with the lawyer to sell the house which left me powerless.” (NI_MYB_DBT_F)*

### Coping Strategies

Findings on NCD outcomes and coping strategies were linked to the Human Model of Disability which frames physical experiences of disease conditions as impairment, particularly if they affected persons due to poverty, stigma and environmental conditions (See figure 1). We have added another dimension to this framing by considering factors such as individual identities, (including poverty and physical manifestations of disease), which in turn shape support systems, and sense of place/belonging and knowledge about disease management as factors shaping the ability to cope with NCDs (See Figure 1)

### Support systems “My wife and children have been paying my bills”

The nexus between NCD-poverty experiences and weak support systems were evident, as people described how they struggled in desperate need for help. NCD participants linked support systems to the nature of help they receive during health shocks from friends and family. Health shocks were described in relation to major physical or health disruptions impacting the ability to seek livelihood. Some participants who sought help from well-wishers said that such support often went towards buying food and medicine and paying rents:

> *“When I was well, I was good at ironing, so I used to help a man to iron his clothes. He has been so good to me, and he is the one who has been paying my house rent.” (NI_CBY_STK_M_01)*

While participants spoke about being in dire need, many felt discouraged about being looked after by others. This was particularly the case when people realised that family members providing care were themselves struggling to meet their own healthcare needs. Participants stated that the nature and length of support they received depended on the level of vulnerability or wellness of their caregivers, some of whom, they said were often stressed about their own health and people in their care as in the case of Ishmail (Box 2). Despite these challenges, caregivers went beyond their limit to support their loved ones with long term health problems:

> *“My wife and children have been paying my bills since I can no longer do business to earn money. So, now I must rely on my family for financial support. However, my wife that has been helping me with the bills has also been suffering from diabetes, chronic cold and myopia. She even had to undergo an eye surgery.” (NI_MYB_DBT_M)*

Where people felt that support from others was running low, stress was often the case. Some participants described having to survive with almost nothing when support from generous people was not forthcoming. Men and women with stroke, widowed and childless women who had limited family support were more likely to be anxious about support drying out:

> *“I was having money from people who were coming to visit me; they helped me financially and with food and medicine, but now they don’t come anymore.” (NI_DZK-HPT-F).*

### Sense of place/tenure status

Insecure tenure was reported as a challenge shaping the experiences of people living with NCDs in informal settlements. Living in one’s own house was tied to how people felt respected within their community. Secure housing tenure was expressed in terms of attachment to a place, and in coping with long-term chronic ill health and social isolation. Participants’ accounts showed that owning a house ignited a sense of comfort and belonging. Where people with a house experienced health shock, such as stroke they said that they were encouraged to cope due to the feeling of respect, due to their secure tenure status:

> *“If I had not built this house, with my condition now, I would have returned to my village. I think the only reason why people respect me in this community is because of my house. Had it not been for that, my situation would have been much worse.” (NI2_DZK_STK_M2)*

However, people living in rented houses stated that insecure housing tenure was a common source of distress, as they felt disrespected, particularly by their landlords. Women’s experiences of housing difficulties reflected the histories of war, forced migration and exacerbated livelihood shocks while living as internally displaced persons in Freetown, causing frequent relocations:

> *“Life has been difficult for me as a single parent…. We have no permanent place to live, which is why we move from one pan-body house (corrugated iron sheet) to another. We lived in several places in Freetown, before we finally moved into this community. The person who rented this place said the land was not his. Therefore, we may be asked to leave at any time the land is requested.” (NI_CBY_DBT_F)*

Women who were widowed or childless were more likely to experience severe housing challenges because of limited access to resources or support. Despite the housing challenges, some people felt connected to their community because of friendships and support they received from others.

### Managing disease symptoms

During health shocks, participants expressed that internalising disease symptoms were key in coping with and managing the physical manifestations of ill health. For many participants, pain, dizziness, fatigue, and blurry vision were the common symptoms that informed them about the intensity of conditions such as hypertension and diabetes. Response strategies included regular physical exercises, medication and adequate rest. Some participants, particularly those with hypertension and stroke spoke about relying on painkillers such as paracetamol to deal with recurring headaches and body pain:

> *“Headache is the main symptom I face, but it usually lessens as soon as I take paracetamol. I also feel severe pain on both of my feet for which I use painkillers." (NI_CBY_HPT_F_01)*

Some participants also described using menthol-based gels, also referred to as ‘hot rubs’ to deal with generalised body pains related to NCDs. It was evident from participants’ framing that deeper understanding about their health helped in not only shaping response to specific symptoms, but in coping with changing environmental circumstances such as weather conditions

> *“I have a hot rub which I apply on the affected part of my body daily. I also buy pain killers from the pharmacy. I feel more pain during the dry season because the intensity of the sun always causes headaches and palpitation of my heart.” (NI_DZK_HPT_M)*

### Drivers of Health Seeking

In this section, our findings show that health seeking decisions by people affected by NCD health conditions are (as shown in figure 1) influenced by three distinct, but related factors: i) Disease condition/trust in provider, ii) Access to healthcare and iii) Health worker attitude.

#### Disease condition/trust in provider

Participants’ type of disease condition and trust in provider influenced how they adapted their health seeking preferences. Across participant groups, different seeking preferences were observed for the treatment of hypertension, disability resulting from stroke, and diabetes. For women with diabetes, the choice of formal healthcare providers was attributed to perceptions of satisfactory outcomes, resulting from perceived correctness and timeliness in the prescription of medicines.

> *“Since I started taking the medicine prescribed from the doctor, I have been feeling better, and I always take it as prescribed” (NI_CBY_DBT_F).*

However, when women with diabetes experienced side effects, slow recovery or recurrent symptoms during biomedical treatment, they were likely to seek traditional remedies such as bitter roots soaked in water, which were believed would reduce their blood sugar. Some women were concerned that side effects related to the constant use certain prescribed drugs caused discomfort as described by Salmata:

> *“I have taken the medicines for a long time, but my body is still deteriorating. I spoke to the doctor, and he advised me to start taking vitamins.” (NI_DZK_DBT_F)*

For women with hypertension, syncretic seeking from formal and informal providers (e.g. drug peddlers) was observed in response to specific symptoms such as headaches and body pain. However, some explained that the lack of improved outcomes from drug peddlers caused them to stick with formal healthcare providers.

Women with hypertension also spoke about traditional remedies such as ginger, garlic and honey which they thought were satisfactory in providing relief from frequent heartbeats. Though with minor side effects such as chest burns, women stated that ginger and garlic were effective in reducing their body weight and “normalising their heartbeats”. Other local remedies such as bitter roots (e.g. “gbangba”) believed to be effective in reducing blood pressure and blood sugar particularly for who reported having the co-morbidities of hypertension and diabetes which one woman summarised as a good remedy that “cleans up the system” (NI_CBY_HPT_F_02).

Women living with stroke, however, spoke about their preference for providers including physiotherapists to enhance their mobility, while preference for drug peddlers was to deal with pain through the purchase of painkillers and ointments. These remedies were sometimes combined with prayers and healing sessions, particularly when recovery from traditional and formal healthcare provider was observed to be slow. Men across NCD categories appeared to have limited interactions with formal healthcare providers. Notably, men with hypertension, stroke and diabetes reported the frequent use of medicines from drug peddlers to deal with generalised body pains associated with their respective NCD conditions. For those who reported visiting the hospital, such visits were commonly dictated by sudden attacks putting them at greater risks of further complications. Men with disability from stroke also said that they sought frequent care from drug peddlers due to their mobility challenges in accessing formal healthcare services. Consequently, some men impacted by stroke expressed preference for drug peddlers due to the availability of drugs and trust in their services:

> *“I buy drugs from a provider called “Wel body na jentri” (Health is wealth) who sells his medicines to community residents. He is very experienced, and his drugs are very effective. He once offered me a card of drugs (10 tablets) when I told him I had no money on that day.” (NI2_CBY_STK_M).*

Men also reported that the use of traditional remedies such as guava, moringa and mango leaves were helpful in dealing with different symptoms of diabetes, as explained by Ishmail (Box 2):

> *“When my feet get swollen, I drink this medicine, and the swelling reduces quickly. It makes me feel better after urinating freely. With the help of ALLAH, my condition is getting better. I don’t remember going to the hospital or pharmacy to buy drugs lately because I trust the traditional medicines that I use.” (NI2_MYB_DBT_M)*

Decision to seek care from formal providers was also influenced by kinship and friendly relationships, particularly those who had medical training. This practice was observed at different points in this study - where participants described how they were introduced either to treatment regimens for different health conditions or warned against care seeking from specific groups of providers such as drug peddlers:

> ***“****I am surrounded by many educated people who always advise me. My nephew is a medical practitioner, and he always warns me against buying drugs from any illegal sources including drug peddlers, as he believes that drug peddlers do not follow correct treatment procedures.” (NI2_MYB_DBT_F)*

#### Healthcare access

Access to healthcare was shaped by the lack of a functioning health centre in the seafront settlement (Cockle Bay), and by geographical impediments in the hillside settlements (Dwarzark and Moyiba).

Because of healthcare access barriers, some participants described that they adapted their health seeking preferences, which included self-replication of treatment based on previously administered treatment by a healthcare provider. Self-replication of treatment, often used by women with diabetes followed the use of previous prescription cards to buy and administer medicines from pharmacies or medicine sellers to treat specific symptoms as described by Salmata *(NI2_DZK_DBT_F)*:

> *“I have already known the medicines to take for diabetes. I usually buy Metformin and Diamine tablets at thCe pharmacy. Through my previous visits to the hospital, I am now familiar with the dosage. When I don’t have money to go to the hospital, I use the prescription to buy the medicines.” (NI2_DZK_DBT_F)*

Men living with hypertension also expressed similar health seeking preferences by seeking home-based treatment from nurses, believing that treatment procedures and outcomes from home-based care were the same as those provided in the hospital.

#### Health worker attitude

Some women who sought care from formal healthcare providers seemed to be generally satisfied with how they were treated. Satisfaction with services from formal providers was characterised as “good treatment” due to the perceived politeness and friendliness of some providers. Other women who preferred buying medicines from the pharmacy noted that providers in the hospital were more concerned about money rather than meeting the needs of patients:

> *“When you go to the hospital, the first thing the doctors will tell you is to be admitted because of the money. The costs of admission, including fees for bed are very high. Hence, I go to the pharmacy. The owner is friendly; he even comes to my house when I am not able to visit the pharmacy.” (NI2_CBY_HPT_F1)*

Experiences about health worker attitude were also expressed in terms of lack of interest and empathy by formal providers. One elderly participant living with disability from stroke recounted his experience during admission at a hospital in Freetown:

> *“The nurses lack empathy and often watch movies on their laptops while patients suffer. They lack total patient care understanding. I was there when a patient in the same ward passed away; no nurse came to his aid to provide him the needed support.” (NI_CBY_STK_M_01)*

However, a few men stated having good experience with formal health providers, including the provision of quality drugs, and treatment on credit, particularly when they had no money:

> *"The doctors have been very kind to me whenever I visit the hospital. They always provide quality drugs and sometimes give me medicines on credit, and I pay back later when I have money.” (NI_MYB_DBT_M)*

## Discussion

This study was undertaken to understand the lived experiences of people affected by NCDs in informal settlements in Freetown. We applied an intersectional analysis to our exploration to consider how disease experience is shaped by different intersecting axes of identity (gender, poverty, disability, place) and amplified by patriarchal disadvantage and livelihood shocks, causing different health outcomes and response capability for men and women. We adapted the Intersectional Gender Analysis Framework for Infectious Diseases of Poverty Research (WHO, 2020) to understand the nexus between NCDs and poverty. The adapted framework provides valuable insights into the theoretical and practical approaches for integrating gender and intersectional analysis into chronic disease research. The framework highlights the complex nature of NCD vulnerabilities.

While efforts are being made within LMIC settings to address gendered social disadvantages such as economic barriers, these considerations are often siloed and do not consider the interactions between economic barriers and other challenges faced by women and men, which shape NCD outcomes (Orozco-Núñez et al., 2024). Within our study, the application of the Intersectional Gender Analysis Framework enabled us to consider the interconnected complexity of social and structural drivers of ill-health shaping the gendered experiences of NCDs in Sierra Leone. As health and social inequities constantly shape negative consequences for marginalised populations in LMICs, an intersectional analysis of poverty, gender and NCD relationships is imperative. Studies have applied these approaches across contexts to understand the different layers of disease vulnerabilities by exploring the connections between Neglected Tropical Diseases (NTDs), disability and mental health (Dean et al., 2022); exploring gender differences and their implications for exposures to infectious disease and outcomes (Tolhurst et al., 2002); analysing stigma related to co-morbidities of obesity, diabetes and hypertension (Rai et al., 2020); and stigma related to HIV, influenced by poverty, gender, sexuality and disability (Turan et al., 2019; Caiola et al., 2014).

Our findings show a strong linkage between poverty, gender stereotypes and the aggravation of NCD disease burden. The poverty-illness nexus was shown through a reduced capacity to respond to crisis (Lumagbas et al., 2018). In the context of NCD vulnerability in informal settlements, entrenched injustice presented through healthcare access barriers, housing difficulties, livelihood shocks and food insecurity were descriptive of the daily crisis which people face in these settings. NCD health problem therefore amplified these pre-existing crises. Women’s lived experiences of NCD conditions reflected historical disadvantage and patriarchal oppression, most notably through their limited financial autonomy, barriers to healthcare decision making and treatment access, compounded by the gendered impacts of conflict and migration. Men’s experiences were however, influenced by changes in social status, due to conflict and migration, and financial instability, limiting access to healthcare. Such differences in gender identities and social barriers presented different outcomes and coping strategies. Our findings are supported by an earlier study that suggests that gendered barriers, including minimal education, lack of financial autonomy, and limited support systems contribute to different capabilities by men and women to respond to ill health (Vlassoff, 2007).

Gender differences, influenced by masculinity were also key in shaping household power dynamics and healthcare decision making. Power dynamics - reflected in the division of roles between men and women - and historical barriers to accessing resources were key factors shaping women’s healthcare decision making (Hawkes et al., 2025). For men living with NCDs, the intersections of masculinity, perceived loss of power, ill health and financial vulnerability were key to how they viewed themselves. Since men played dominant roles in household decision making, financial collapse and failing health signalled “loss of power” through diminished decision-making power. This was often a source of marital conflict, impacting men’s ability to cope with NCD health problems.

Within the global health system, the framing of masculinity has often tended to focus on power dynamics and the social disadvantages faced by women, without adequately exploring how it also negatively affects men (Hawkes et al., 2025). Therefore, achieving global health policies and interventions that prioritise gender equity to improve population health outcomes remains a challenge (Hawkes et al., 2025). Policies with limited focus on masculinity have the potential to limit men’s access to healthcare. Chikovore et al., (2020) cited stigma and the feeling of not being welcomed as factors that inhibit men from seeking care at primary healthcare facilities for conditions such as tuberculosis (TB) and HIV - as these facilities are believed to prioritise maternal and child health issues. Similarly, worse health outcomes have been identified among men living with NCD conditions because of limited or delayed access to healthcare (Ngaruiya, 2022). Similar patterns were observed in this study as men with hypertension, diabetes and disability resulting from stroke were more inclined to interact with drug peddlers due to barriers to formal healthcare services, shaped by financial challenges, distance and disability.

Coping with the burden of NCD conditions in informal settlements is undermined by limited support systems and the lack of services for NCDs in informal settlements. Healthcare access barriers faced by people living with NCD, combined with daily challenges (e.g. poverty, livelihood shocks, and housing insecurity) limit their physical and social functioning, often leading to disability. The Human Development Model of Disability (Mitra, 2017) describes disability resulting from health conditions in terms of deprivation of support in enhancing capability and functioning.

The model of disability aligns with our findings, as people living with NCDs sometimes felt disabled due to deprivation in meeting their social and healthcare needs. Therefore, people often adapted different strategies to cope with the burden of NCDs. Men with secure housing tenure, social networks and family support had better advantage over women in coping with the burden of NCD. People with limited social support - such as widows and childless women faced more difficulty in coping with the burden of NCDs due to insecure housing and the feeling of stigma. This requires health systems to adapt to the complex drivers shaping NCD vulnerabilities by designing context specific healthcare interventions.

The strength of this study is the application of a gender and intersectional lens to the study of NCDs in urban informal settlements. The application of the Gender and Intersectional Analysis of Infectious Disease of Poverty Research (WHO, 2020) in a new context (i.e. in informal settlements in Freetown) has enhanced our understanding of the disease-poverty nexus. An important contribution is the application of this framework specifically to NCDs, which brings a new understanding of NCD interaction with poverty and gender inequities.

By building on the framework’s previous application to infectious diseases, its adaptation to NCDs in Freetown has provided us with additional insights on how structural inequities worsen disease outcomes. One of the leanings drawn from this study is that NCDs impact women and men differently which means that the delivery of healthcare services must be designed in a people centered approach that integrates the perspectives of patients and providers for an effective management of long-term illness (United Nations, 2015). Finally, health systems must be gender responsive to ensure that the healthcare needs of patients with diverse social identities are adequately addressed.

One of the limitations of this study is its limited geographic scope - being conducted in urban informal settlements - which excludes the diverse experiences of other urban populations living with NCDs in Freetown. However, conducting this study in urban informal settlements helps address evidence gaps on NCDs in complex and marginalised urban contexts. Future studies should explore the voices of urban residents across varying layers of vulnerability to understand how these vulnerabilities are shaped.

## Conclusion

Findings from this study reveal strong linkages between poverty, gender inequality and disability in shaping the burden of NCDs. Gender norms and patriarchal disadvantages reinforce power hierarchies and exacerbate poverty and negative health outcomes. Health systems should adapt to the challenges that reinforce the burden of NCDs in rapidly changing urban environments (e.g. poverty and social inequalities) by ensuring that healthcare delivery meets the needs of patients with diverse social identities. This aligns with our findings which reveal that women’s lived experiences of NCDs reflected gendered historical disadvantages (e.g. patriarchal oppression) that limit financial autonomy and healthcare access by women while, men’s experiences were influenced by changes in social status, due to conflict and migration, and financial instability, limiting their access to healthcare. This requires that health systems should design gender responsive strategies to address NCD burdens, particularly in marginalised urban settings.

## Data Availability

All data in this study are contained in this manuscript

## Notes

Drug peddlers in Sierra Leone are informal healthcare providers known for selling medicines to clients particularly in areas healthcare services are not easily accessed like in informal settlements. Many of these providers are unlicensed.

Pan-body houses are low-cost houses built typically in informal settlements using corrugated iron sheets and sticks as wells and for roofing. These houses are used as temporary or permanent housing solutions for low-income earners. They are exposed to risks due to the poor-quality materials.

## Authors Contributions

AC conceptualised research ideas analysed data and wrote the first draft; LD, ST & AW conceptualised research ideas, interpreted data and reviewed first draft; JM & BK interpreted date and reviewed final draft. All authors reviewed and approved the final draft.

## Acknowledgements

We are grateful to all co-researchers living and working in research sites and Research Assistants for helping with participant recruitment within their communities for conducting some of the key informant interviews.

## Ethical Approval

The ethics application for this study was approved by the Sierra Leone Ethics and Scientific Review Committee (Version number 2.0 of 28^th^ July 2021) on 3rd August 2021 after fulfilling all requirements. Ethics application was also approved by the Research and Ethics Committee of the Liverpool School of Tropical Medicine (LSTM) (Version number 21-043) on 5th August 2021. Both approval bodies requested the submission of all research tools including the narrative interview guide, information sheet, and consent form. The ethics approvals provided adequate safeguards for the protection of research participants and the research team from harm.

